# Field Testing of a Decision Support Tool for Acute Appendicitis using an Online Randomized Experimental Design

**DOI:** 10.1101/2021.11.09.21265975

**Authors:** Joshua E. Rosen, David R. Flum, Giana H. Davidson, Joshua M. Liao

## Abstract

**Background:** Mounting evidence from randomized controlled trials have shown that antibiotics can be a safe and effective treatment for appendicitis. Patients and surgeons must work together to choose the optimal treatment approach for each patient based on their own preferences and values. We developed a novel decision support tool (DST) to facilitate shared decision making for appendicitis. The effect of this DST on decisional outcomes remains unknown.

**Methods:** We conducted an online randomized field test in at-risk individuals comparing the DST to a standard infographic as a control. Individuals were randomized 3:1 to the DST (intervention) or infographic (control). The primary outcome was the total decisional conflict scale (DCS) score measured before and after exposure to the DST. Secondary outcomes included between-group DCS scores, and between group comparisons of the acceptability of DST or infographic.

**Results:** 180 individuals were included in the study after quality control checks. Total DCS scores decreased significantly after viewing the DST (59.0 to 14.6, p<0.001) representing movement from a state of high to low decisional conflict. Individuals exposed to the DST reported higher acceptability ratings (3.7 vs. 3.3, p<0.001) and had more individuals who would completely agreed that they would be willing to try antibiotics (45% vs. 21%, p=0.008).

**Conclusions:** The novel appendicitis DST significantly decreased decisional conflict in this online randomized field test. Users rated the tool as highly acceptable and expressed increased willingness to consider antibiotics as a treatment approach after viewing it.

**Study Funding:** This work was supported by the National Institute of Diabetes and Digestive and Kidney Diseases [grant number T32DK070555]; and a generous gift from Marty and Linda Ellison. The development of aspects of the decision support tool was funded by the Patient Centered Outcomes Research Institute. The funding sources had no role in the study design, collection, analysis, or interpretation of data, in the writing of the manuscript or in the decision to submit for publication.

## Introduction

Appendicitis is a common acute condition typically treated with surgery (appendectomy). Recently, several randomized controlled trials have demonstrated the safety and efficacy of using antibiotics to treat appendicitis.(1–5) In particular, the Comparison of Outcomes of Antibiotic Drugs and Appendectomy (CODA) trial found similar outcomes for the two treatments in terms of patient-reported quality of life at 30 and 90 days, and similar time to symptom resolution. However, surgery and antibiotics differ with respect to many outcomes that may be important for patients, such as the risk of needing future procedures, number of days of work missed, and risk of occult malignancy. Furthermore, patients and clinicians may prioritize outcomes differently.(6) As a result, appendicitis treatment decisions are both highly preference-sensitive and ripe for strong shared decision making between patients and clinicians.

Given these factors, as well as the time-pressure frequently surrounding treatment decisions (i.e., in the emergency department between patients and clinicians without long-standing relationships), there is a need for better tools to support informed shared decision making. Unfortunately, few such resources exist. Therefore, using the results from the CODA trial, we developed an appendicitis decision support tool (DST) (www.appyornot.org). The DST consists of an informational video and an interactive decision aid to help patients with appendicitis understand differences in outcomes between surgery and antibiotics, and pick the one that best aligns with their preferences and values.

Here we describe the results of an online randomized field test of the DST versus a standard infographic. The goals of the field test were to assess the effect of the DST on decisional outcomes among an at-risk population of US adults.

## Methods

### Study Population

We utilized Amazon’s Mechanical Turk (MTurk) crowdsourcing platform to test the DST in a simulated at-risk population of American adults (≥ 18 years old). Participants did not actively have appendicitis but represented an at-risk population since appendicitis is one of the most common acute surgical conditions in the United States, and the typical age range of participants in our group’s prior MTurk studies(6,7) has matched participants from the CODA trial. Participants were required to have an MTurk approval rating ≥ 95% and were excluded from the study if they had previously had appendicitis or were employees of the University of Washington. Participants provided demographic information during the survey and completed an objective numeracy scale.(8) The University of Washington institutional review board considered this study exempt. Individuals were paid $5 for participation.

### Description of Intervention and Control

Participants were randomized in a 3:1 ratio to an intervention versus control arm, respectively. In the intervention arm, participants were exposed to the DST consisting of both an informational video and interactive patient decision aid. In the control arm, participants were exposed to a previously published infographic describing the results of the CODA trial (http://becertain.org/coda-infographic), which provided basic information, but not the detailed discussions, comparisons, and value-clarification activities present in the DST. Since participants were not directly interacting with medical providers, we could not use “usual clinical care” for the control arm.

### Outcomes

The primary outcome was the total score on the decisional conflict scale (DCS) in a pre-post analysis in the intervention arm. The DCS (9,10) is a validated instrument for measuring decisional conflict and has been used in numerous studies of decision-support interventions.(11) The DCS has several subscales that speak to different dimensions of the decision-making process that may contribute to decisional conflict including: feeling *informed, values clarity*, feeling *uncertain*, feeling *supported*, and making an *effective decision*. We considered these to be explanatory factors of the primary outcome that provide additional detail in order to understand the dimensions of decisional conflict that the intervention may effect. The DCS and its subscales are scored on a 100-point scale with lower scores representing less decisional conflict. Scores over 37.5 are typically considered to reflect very high decisional conflict associated with feeling unsure.(10,12) Secondary outcomes included between-arm comparisons of the DCS, as well as between-arm comparisons of decision aid acceptability (measured on a 4-point scale) and perceptions of trust and accuracy in the information (5-point scale). To remove questions not applicable to our online field test design, we used a modified version of the DCS that excluded the *Support* subscale and a question about “sticking with my decision” from the *Effective Decision* subscale. Scores were re-scaled appropriately on the 100-point scale as per instructions in the DCS User Manual.(10)

### Study Quality Control

We implemented several quality control methods including a Completely Automated Public Turing Test to Tell Computers and Humans Apart (CAPTCHA) test, an attention-check question at the end of the survey, and manual review of survey responses for duplicate MTurk IDs. Furthermore, to ensure that participants viewed the entire DST, we required participants in the intervention arm to enter a passcode displayed at the end of the DST to advance in the survey.

## Results

Overall, 194 individuals completed the survey and passed quality control checks. 14 participants reported or were unsure about a prior diagnosis of appendicitis and were excluded from the analysis (1 in the control arm, 13 in the intervention arm). Among 180 participants in the final analysis, demographic characteristics were well-balanced between study arms (Table 1). Prior to participating in this study, the majority of individuals in both arms knew that appendicitis was commonly treated with surgery (87.5% in control arm, 90.2% in intervention arm, p=0.81) and the minority were aware that antibiotics were an option to treat appendicitis (12.5% in control arm, 6.1% in intervention arm, p=0.27).

**Table 1:** Participant Characteristics.

| Variable | Level | Control<br>n = 48 | Intervention<br>n = 132 | p-value |
| --- | --- | --- | --- | --- |
| Age (Mean(SD)) |  | 38.46 (10.41) | 40.40 (11.80) | 0.316 |
| Sex |  |  |  | 0.214 |
|  | Female | 17 (35.4) | 54 (40.9) |  |
|  | Male | 30 (62.5) | 78 (59.1) |  |
|  | Prefer not to say | 1 (2.1) | 0 (0.0) |  |
| Gender Identity |  |  |  | 0.214 |
|  | Female | 17 (35.4) | 54 (40.9) |  |
|  | Male | 30 (62.5) | 78 (59.1) |  |
|  | Other | 1 (2.1) | 0 (0.0) |  |
| Racial Identity |  |  |  | 0.675 |
|  | Black/African American | 3 (6.2) | 10 (7.6) |  |
|  | East Asian | 3 (6.2) | 8 (6.1) |  |
|  | Multiple identities | 2 (4.2) | 5 (3.8) |  |
|  | Other | 0 (0.0) | 3 (2.3) |  |
|  | South Asian | 1 (2.1) | 2 (1.5) |  |
|  | Unknown | 1 (2.1) | 0 (0.0) |  |
|  | White | 38 (79.2) | 104 (78.8) |  |
| Ethnicity |  |  |  | 0.58 |
|  | Hispanic/Latino/Latinx | 3 (6.2) | 5 (3.8) |  |
|  | Non-Hispanic/Latino/Latinx | 44 (91.7) | 126 (95.5) |  |
|  | Prefer not to say | 1 (2.1) | 1 (0.8) |  |
| Education Level |  |  |  | 0.361 |
|  | 2-year College Degree | 4 (8.3) | 13 (9.8) |  |
|  | 4-year College Degree | 21 (43.8) | 57 (43.2) |  |
|  | Graduate Degree | 7 (14.6) | 9 (6.8) |  |
|  | High School / GED | 6 (12.5) | 23 (17.4) |  |
|  | Some College | 9 (18.8) | 28 (21.2) |  |
|  | Some High School | 0 (0.0) | 2 (1.5) |  |
|  | Unknown | 1 (2.1) | 0 (0.0) |  |
| Insurance |  |  |  | 0.606 |
|  | Employer-Provided Insurance | 25 (52.1) | 56 (42.4) |  |
|  | Government | 2 (4.2) | 3 (2.3) |  |
|  | MediCAID | 5 (10.4) | 17 (12.9) |  |
|  | MediCARE | 2 (4.2) | 10 (7.6) |  |
|  | Not Insured | 8 (16.7) | 18 (13.6) |  |
|  | Other | 2 (4.2) | 4 (3.0) |  |
|  | Private Insurance | 4 (8.3) | 24 (18.2) |  |
| Employment Status |  |  |  | 0.144 |
|  | Employed full-time | 32 (66.7) | 84 (63.6) |  |
|  | Employed part-time | 6 (12.5) | 17 (12.9) |  |
|  | Prefer not to say | 1 (2.1) | 0 (0.0) |  |
|  | Retired | 0 (0.0) | 5 (3.8) |  |
|  | Self-employed | 6 (12.5) | 21 (15.9) |  |
|  | Student | 2 (4.2) | 0 (0.0) |  |
|  | Unemployed (looking for work) | 1 (2.1) | 4 (3.0) |  |
|  | Unemployed (not looking for work) | 0 (0.0) | 1 (0.8) |  |
| Annual Household Income |  |  |  | 0.385 |
| | <\$25,000 | 8 (16.7) | 20 (15.2) | |
| | >\$100,000 | 9 (18.8) | 12 (9.1) | |
| | \$25,000 - \$50,000 | 12 (25.0) | 43 (32.6) | |
| | \$50,001 - \$75,000 | 13 (27.1) | 33 (25.0) | |
| | \$75,001 - \$100,000 | 5 (10.4) | 23 (17.4) | |
|  | Prefer not to say | 1 (2.1) | 1 (0.8) |  |
| Before taking this survey,<br>did you know that<br>appendicitis is commonly<br>treated with surgery? |  | 42 (87.5) | 119 (90.2) | 0.812 |
| Before taking this survey,<br>did you know that it is<br>possible to treat appendicitis<br>with antibiotics? |  | 6 (12.5) | 8 (6.1) | 0.266 |
| Objective Numeracy (mean<br>(SD)), max score = 3 |  | 2.38 (0.87) | 2.33 (0.81) | 0.725 |

DCS scores decreased after viewing the DST (Table 2), reflecting a movement from a very high to a low state of decisional conflict.(10) In particular, average DCS total scores decreased from 59.0 to 14.6 (p<0.001). DCS scores significantly decreased across all subscales (Table 2) indicating that the DST effectively addressed multiple areas of decisional conflict. Participants also scored higher on an objective knowledge test of information relevant to the decision after viewing the DST (3.4 vs. 2.0 out of 5, p<0.001).

**Table 2:** Pre vs. Post exposure comparisons in the group who viewed the decision support tool.

|  | Pre-Exposure to DST<br>(Mean (SD)) | Post-Exposure to DST<br>(Mean (SD)) | p-value |
| --- | --- | --- | --- |
| Knowledge | 1.98 (1.01) | 3.42 (0.76) | <0.001 |
| DCS Total | 59.06 (22.60) | 14.57 (13.06) | <0.001 |
| DCS Informed<br>Subscale | 52.21 (18.90) | 11.43 (11.67) | <0.001 |
| DCS Values Clarity<br>Subscale | 60.23 (27.58) | 13.32 (13.69) | <0.001 |
| DCS Uncertainty<br>Subscale | 67.11 (27.19) | 19.76 (19.74) | <0.001 |
| DCS Effective<br>Decision Subscale | 56.69 (26.04) | 13.76 (14.38) | <0.001 |

\*DCS = Decisional Conflict Scale

There was no significant difference in baseline DCS or knowledge scores between the intervention or control arms. After exposure to the DST or control infographic, mean acceptability ratings (3.7 vs. 3.3 out of 4, p<0.001) as well as perceived trust in (4.5 vs. 4.3 out of 5, p=0.02) and perceived accuracy of (4.7 vs. 4.4 out of 5, p=0.005) the information was significantly higher for the DST. Total DCS and subscale scores were lower for the DST compared to the control (Table 3), but this difference was not statistically significant except for the effective decision subscale (13.8 vs 20.1, p=0.02). Notably, the range of effect sizes (defined as the mean difference divided by the pooled standard deviation) range from 0.09 to 0.39 for the DCS and its subscales – values that were in the range of those typically seen in formal randomized trials of decision support interventions.(10,11)

**Table 3:**
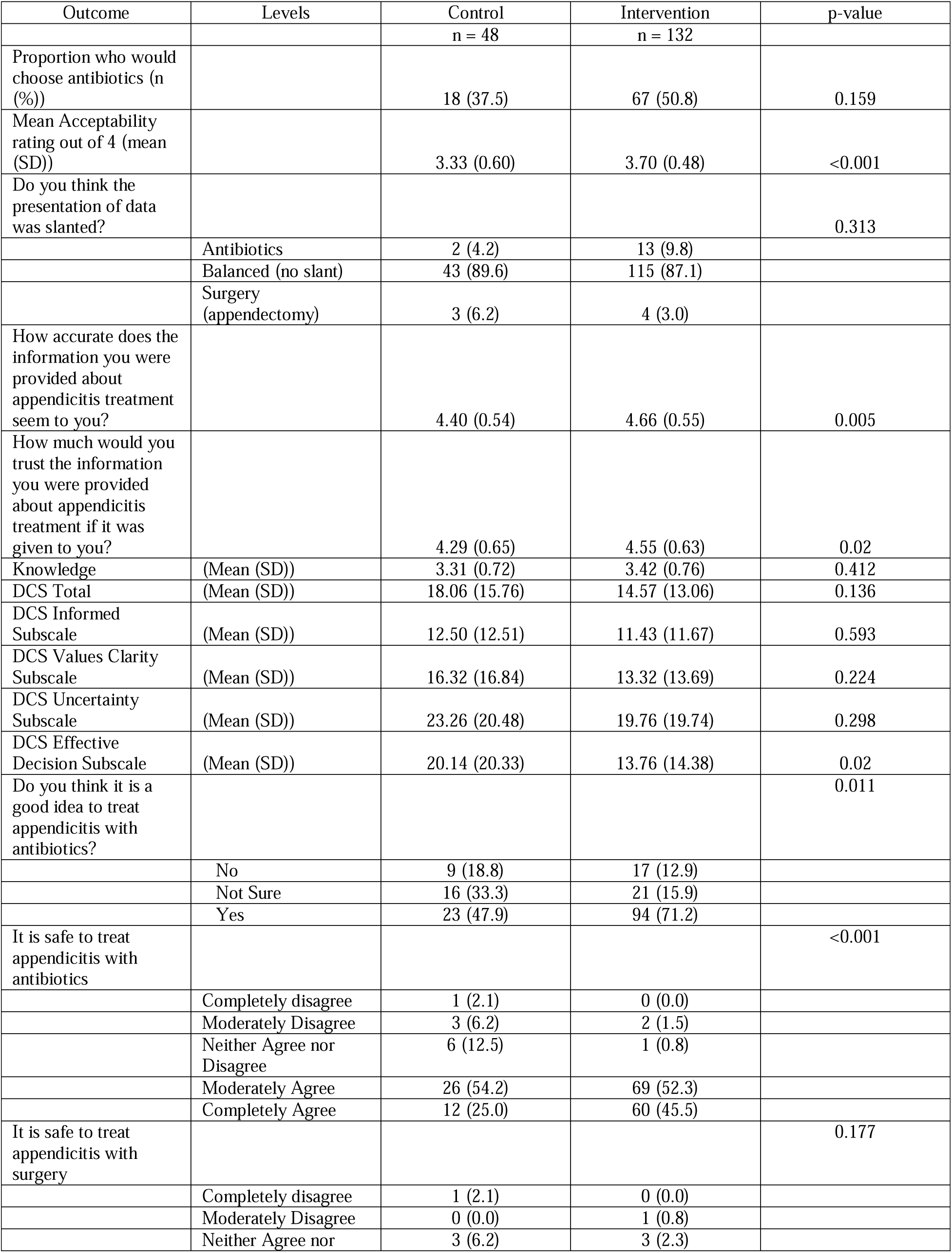
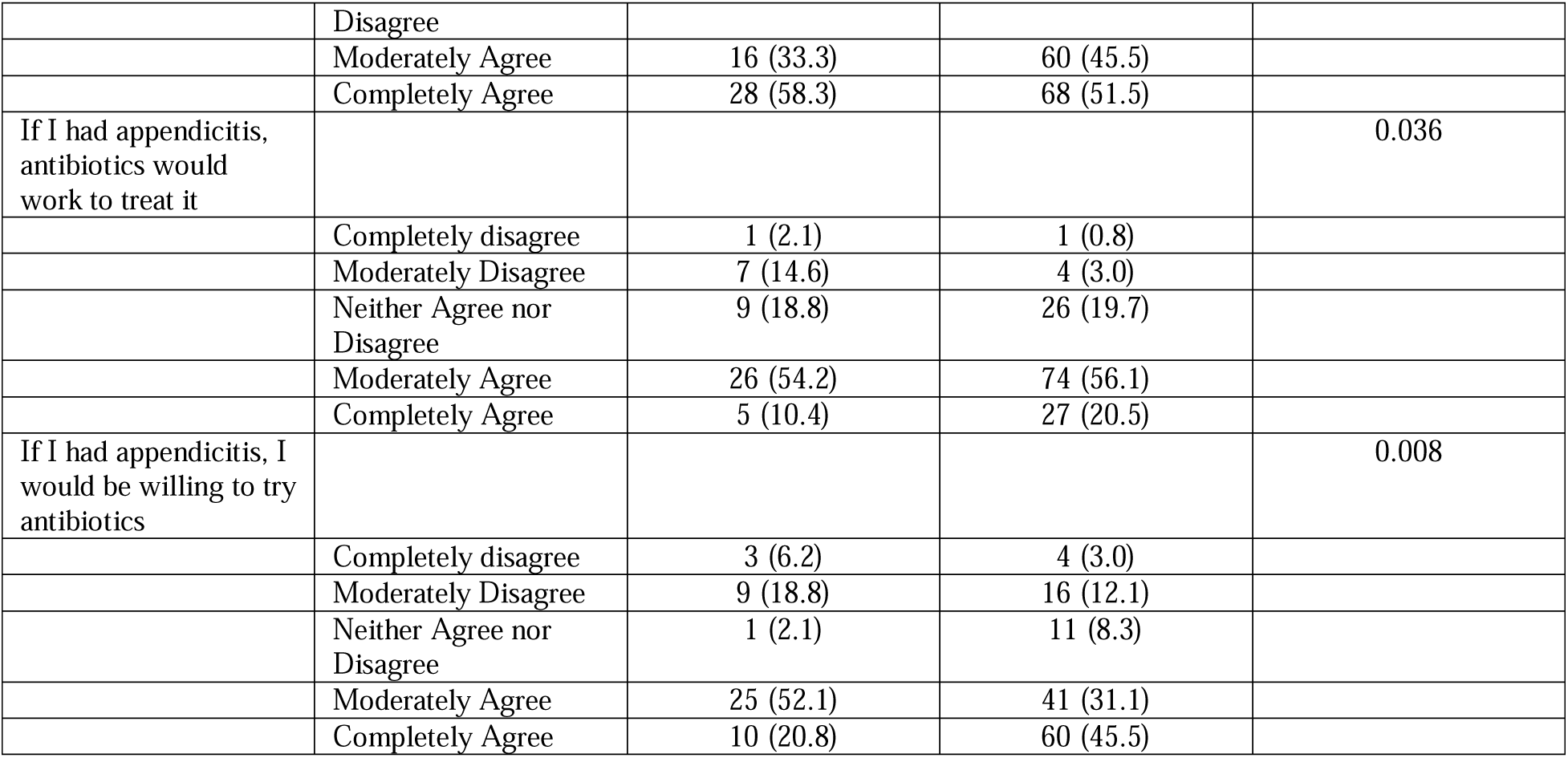
Between Arm Comparisons of post-exposure outcomes.

Participants were asked a series of questions about their opinions regarding the use of antibiotics and surgery for treating appendicitis after viewing either the DST or control infographic (Table 3). Significantly more participants in the intervention arm thought that it was a good idea to treat appendicitis with antibiotics (71% vs 48%, p=0.011) and agreed that it was safe to treat appendicitis with antibiotics (97% vs. 79%, p<0.001), while similar proportions agreed that it was safety to treat appendicitis with surgery (97% vs. 92%, p=0.18). Participants exposed to the DST also had stronger agreement that antibiotics would work for them if they had appendicitis (77% vs. 65%, p=0.036) and indicated a willingness to try antibiotics (77% vs. 73%, p=0.008). While more participants in the DST arm stated they would choose antibiotics to treat appendicitis in the proposed scenario, this difference was not statistically significant (50.8% vs. 37.5%, p=0.16)

## Discussion

This paper reports the results of an online field-test of a novel DST for acute appendicitis. It shows that compared to a standard infographic, the DST significantly decreased decisional conflict and achieved higher acceptability, trust, and accuracy ratings. Several points are notable.

First, the DST achieved high acceptability ratings compared to the infographic, while requiring much more time to view and providing more complex and nuanced numerical information. These are strong preliminary signals about feasibility that suggest patients could find the DST usable in a clinical environment, and in turn support future clinical testing.

Second, the DST moved DCS scores by a large magnitude in the intended direction, from a region of high decisional conflict to low decisional conflict. In particular, the scores for the *values-clarity* and *informed* DCS subscales were very low after viewing the DST, indicating its effectiveness in these two key areas. These effects are particularly notable given differences between this simulated environment and real clinical environments, in which individuals would view the DST in the context of discussions with a healthcare provider. In the absence of that context, our findings highlight the DST as a potentially effective intervention for improving decisional outcomes even in settings where surgical consultation and detailed in-person discussion may not immediately available. Though differences were not statistically significant, the DCS was lower in the intervention versus control arm and effect sizes were in the range of those often seen in studies of DSTs.(10,11)

Compared to those viewing the control infographic, participants viewing the DST indicated stronger agreement that antibiotics are a safe option for treating appendicitis. This finding is particularly salient given that like populations from other studies, individuals in our sample recognized surgery as the widely accepted treatment for appendicitis. Thus, the DST served as an effective intervention to inform patients and increase their comfort about antibiotics a new alternative. Notably, we observed this effect for the DST without concomitant perceptions that the tool biased decisions toward either surgery or antibiotics. It is possible that the DST increased trust and accuracy ratings among participants, which in turn increased their willingness to consider an alternative treatment option.

Study limitations include sample size, at-risk but not actively ill population, and use of an infographic in place of usual clinical care in the control arm. The infographic may have biased toward null difference between arms given that it included more information than typically provided in many clinical interactions. This study was also not designed or powered to detect differences of this size between study arms.

In conclusion, this new DST for acute appendicitis decreased decisional conflict, was highly acceptable to users, and encouraged consideration of antibiotics as a new treatment approach. These results are promising and support the testing of this novel DST and its effect on clinical decision making among patients suffering from acute appendicitis.

## Data Availability

All data produced in the present study are available upon reasonable request to the authors

## References

1. Collaborative C, Flum DR, Davidson GH, Monsell SE, Shapiro NI, Odom SR, et al. A Randomized Trial Comparing Antibiotics with Appendectomy for Appendicitis. New Engl J Med. 2020;383(20):1907–19.

2. O’Leary DP, Walsh SM, Bolger J, Baban C, Humphreys H, O’Grady S, et al. A Randomised Clinical Trial Evaluating the Efficacy and Quality of Life of Antibiotic Only Treatment of Acute Uncomplicated Appendicitis: Results of the COMMA trial. Ann Surg. 2021;Publish Ahead of Print.

3. Salminen P, Tuominen R, Paajanen H, Rautio T, Nordström P, Aarnio M, et al. Five-Year Follow-up of Antibiotic Therapy for Uncomplicated Acute Appendicitis in the APPAC Randomized Clinical Trial. Jama. 2018;320(12):1259–65.

4. Sallinen V, Akl EA, You JJ, Agarwal A, Shoucair S, Vandvik PO, et al. Meta□analysis of antibiotics versus appendicectomy for non□perforated acute appendicitis. Brit J Surg. 2016;103(6):656–67.

5. Park HC, Kim MJ, Lee BH. Randomized clinical trial of antibiotic therapy for uncomplicated appendicitis. Brit J Surg. 2017;104(13):1785–90.

6. Rosen JE, Agrawal N, Flum DR, Liao JM. Willingness to undergo antibiotic treatment of acute appendicitis based on risk of treatment failure. Brit J Surg. 2021;znab280-.

7. Rosen JE, Agrawal N, Flum DR, Liao JM. Verbal Descriptions of the Probability of Treatment Complications Lead to High Variability in Risk Perceptions: A Survey Study. Ann Surg. 2021;Publish Ahead of Print.

8. Schwartz LM, Woloshin S, Black WC, Welch HG. The Role of Numeracy in Understanding the Benefit of Screening Mammography. Ann Intern Med. 1997;127(11):966.

9. O’Connor AM. Validation of a Decisional Conflict Scale. Med Decis Making. 1995;15(1):25–30.

10. O’Connor A. User Manual - Decisional Conflict Scale (16 item statement format) [Internet]. 2010 [cited 2021 Oct 19]. Available from: http://decisionaid.ohri.ca/docs/develop/User_Manuals/UM_Decisional_Conflict.pdf

11. Stacey D, Légaré F, Lewis K, Barry MJ, Bennett CL, Eden KB, et al. Decision aids for people facing health treatment or screening decisions. Cochrane Db Syst Rev. 2017;4(4):CD001431.

12. Kryworuchko J, Stacey D, Bennett C, Graham ID. Appraisal of primary outcome measures used in trials of patient decision support. Patient Educ Couns. 2008;73(3):497–503.

