## Supplementary Material 1 for "Field Testing of a Decision Support Tool for Acute Appendicitis using an Online Randomized Experimental Design"

**Supplementary Material – AAPOR Reporting Guidelines**

| **Checklist Item** | **Response** |
| --- | --- |
| Survey Sponsor | University of Washington |
| Survey/Data Collection Supplier | Survey created on Qualtrics. Survey administered on Amazon’s Mechanical Turk platform via the CloudResearch Inc. interface |
| Population represented | Adults in the United States |
| Sample Size | 194 -> 180 complete respondents |
| Mode of Data Collection | Internet survey |
| Type of sample | Non-probability |
| Start and end dates of Data Collection | 10/6/2021 |
| Margin of sampling error for total sample | N/A |
| Are the data weighted? | No |
| Subgroup reporting | No |
